# Impact of ineffective prehospital analgesia on chronic pain outcomes. A secondary analysis of outcomes from the PACKMaN randomised controlled trial

**DOI:** 10.1101/2025.08.01.25332769

**Authors:** Michael A Smyth, Hannah Noordali, Kath Starr, Joyce Yeung, Ranjit Lall, Felix Michelet, Gordon Fuller, Stavros Petrou, Alison Walker, Zoe Green, Rebecca McLaren, Elisha Miller, Gavin D Perkins

## Abstract

**Introduction:** Previous research establishes that patients often experience inadequate pain relief following acute traumatic injury. The aim of this secondary analysis of data from the PACKMaN trial is to explore the contributing factors and impact of sub-optimal pain relief on short to long term outcomes.

**Methods:** PACKMaN was a randomised controlled trial comparing ketamine and morphine when used by paramedics treating severe pain following trauma. We dicotomised all patients into two groups, those who reported moderate to severe pain (NRS pain score ≥ 4) at hospital arrival and those who reported mild pain at hospital arrival (NRS pain score < four), irrespective of which treatment they had received. We explore the contributing factors to analgesic efficacy, hospital, short (3 month) and long-term (6 month) pain outcomes.

**Results:** Final pain score was available in 446 (99%) participants. In our analysis population 175 (39%) had a final pain score below 4 and 271 (61%) had a final pain score of 4 or above. Significant differences were found in the Sum of Pain Intensity Difference (SPID), Total Pain Relief (TOTPAR) scores, percentage of maximum dose in milligrams, patient global impression of change, time to perceptible analgesia, time to meaningful analgesia, and duration of analgesia, participants being admitted to hospital in the adjusted analysis, and both short and long-term pain outcomes on the brief pain inventory.

**Conclusion:** Participants still experiencing moderate to severe pain at hospital arrival report greater pain severity scores at both 3- and 6-months post randomisation as well as higher pain interference scores.

**Trial Registration:** ISRCTN14124474

## Introduction

It has been reported that up to 24% of ambulance calls are related to trauma.(1) Published data indicate that pain is present in at least 70% of trauma patients attended by ambulance services.(2) Furthermore, where a pain score is documented, 81% of patients report moderate or severe pain.(2)

Pain may precipitate several adverse physiological effects including haemodynamic,(3–5) respiratory(6) and metabolic(7) compromise. In addition, the World Health Organisation (WHO) assert that effective pain management should be a basic human right.(8) Pain management is a therefore key priority for ambulance clinicians.(9).

Evidence suggests pain is poorly managed within ambulance services.(10–14) Poorly managed acute pain is associated with adverse patient outcomes(15) including depression, post-traumatic stress disorder (PTSD) and chronic pain.(16) Chronic pain is defined as pain that persists, or recurs, for more than 3 months,(17) and it is associated with significant economic and social burden.(18–20)

A direct relationship has been established between acute pain severity and chronic pain.(16) Studies have demonstrated that effective treatment of acute pain reduces the risk of developing chronic pain.(21, 22). Previous research has investigated the prevalence of chronic pain following surgery, (23–26) but few studies have explored the impact of prehospital pain management on the prevalence of chronic pain and other important clinical outcomes.

The Paramedic Analgesia Comparing Ketamine and MorphiNe (PACKMaN) trial was a multi-centre, double-blinded randomised controlled trial (RCT) comparing the clinical and cost effectiveness of ketamine and morphine for severe pain following acute traumatic injury (EudraCT 2020-000154-10, ISRCTN14124474). The trial protocol(27) and results(28) have previously been published.

This secondary analysis explores the difference in outcomes reported in the PACKMaN trial, dichotomised by pain severity on arrival at hospital. We compared patients whose pain was successfully reduced from severe (NRS≥7/10) to mild (NRS < 4/10), and those who were still experiencing moderate or severe pain (NRS≥4/10) upon arrival at hospital, irrespective of which trial drug they received.

## Methods

### Setting

PACKMaN was undertaken in two English Ambulance Services – West Midlands Ambulance University NHS Foundation Trust (WMAS) and Yorkshire Ambulance Service NHS Trust (YAS). WMAS serves a population of 5.6 million over a geographical area of 5500 square miles (WMAS, 2024) while YAS serve 5 million people over 6000 square miles (YAS,2024). Collectively, these services receive more than 2 million 999 emergency calls annually and employs more than 14,000 staff.

### Participants

Patients were eligible for inclusion if they had suffered an acute traumatic injury, were at least 16 years of age or over, reported a pain score of 7 or greater on a 0-10 numeric rating score (NRS) and, in the opinion of the attending paramedic, would normally require parenteral morphine for analgesia.

Patients were excluded if they were known or suspected to be pregnant, unable to articulate severity of pain using the 0-10 NRS, lacked capacity due to a reason other than pain, were a prisoner or they declined participation. In addition, patients who received parenteral ketamine or opioid analgesia prior to randomisation, or who had a contraindication to either drug, were also excluded.

### Ethical approval

Ethical approval was provided by the West of Scotland Research Ethics Committee (20/WS/0126).

### Consent

A delayed consent model was approved by the ethics committee. The attending paramedic advised patients meeting the eligibility that they would like to enrol them into the PACKMaN trial. Brief information about the trial was provided and verbal assent to participation was sought by the paramedic prior to randomisation. Informed, written consent was obtained at a later stage by trained research paramedics, either during the patient’s hospital stay, or following their discharge from hospital. If the patient declined participation standard care was provided.

### Intervention

Patients were randomised to receive either morphine sulphate (control) or ketamine hydrochloride (intervention). Drugs were administered intravenously and titrated to effect. The maximum cumulative dose of morphine was 20mg, while the maximum cumulative dose of ketamine was 30mg.

### Randomisation

To ensure allocation concealment, trial drugs were packaged in sequentially numbered treatment packs adhering to a permuted, unstratified, block randomisation system (variable size blocks) providing an overall ratio of 1:1 control: intervention. The randomisation sequence was generated by the trial statistician and randomisation occurred when the paramedic opened the drug-pack. Only the trial statistician was able to access the unblinded data. All other participants, including paramedics and patients, were blinded to the treatment delivered.

### Variables

In PACKMaN, the primary outcome was the Sum of Pain Intensity Difference (SPID) score.(29) SPID is defined as difference in pain intensity over the assessment period.(23) Pain intensity was quantified using the numeric rating scale (NRS). The NRS is a validated tool with scores ranging from 0-10, with 0 representing no pain and 10 representing the worst pain imaginable.(30) It can be further used to categorise pain - those with a score between 0 and 3 are said to have mild pain, while a NRS score between 4 and 6 are said to have moderate pain, and a NRS score of 7 or above is said to represent severe pain. (24) Secondary outcomes, including the time points at which they were measured, are reported in Table 1.

**Table 1.**
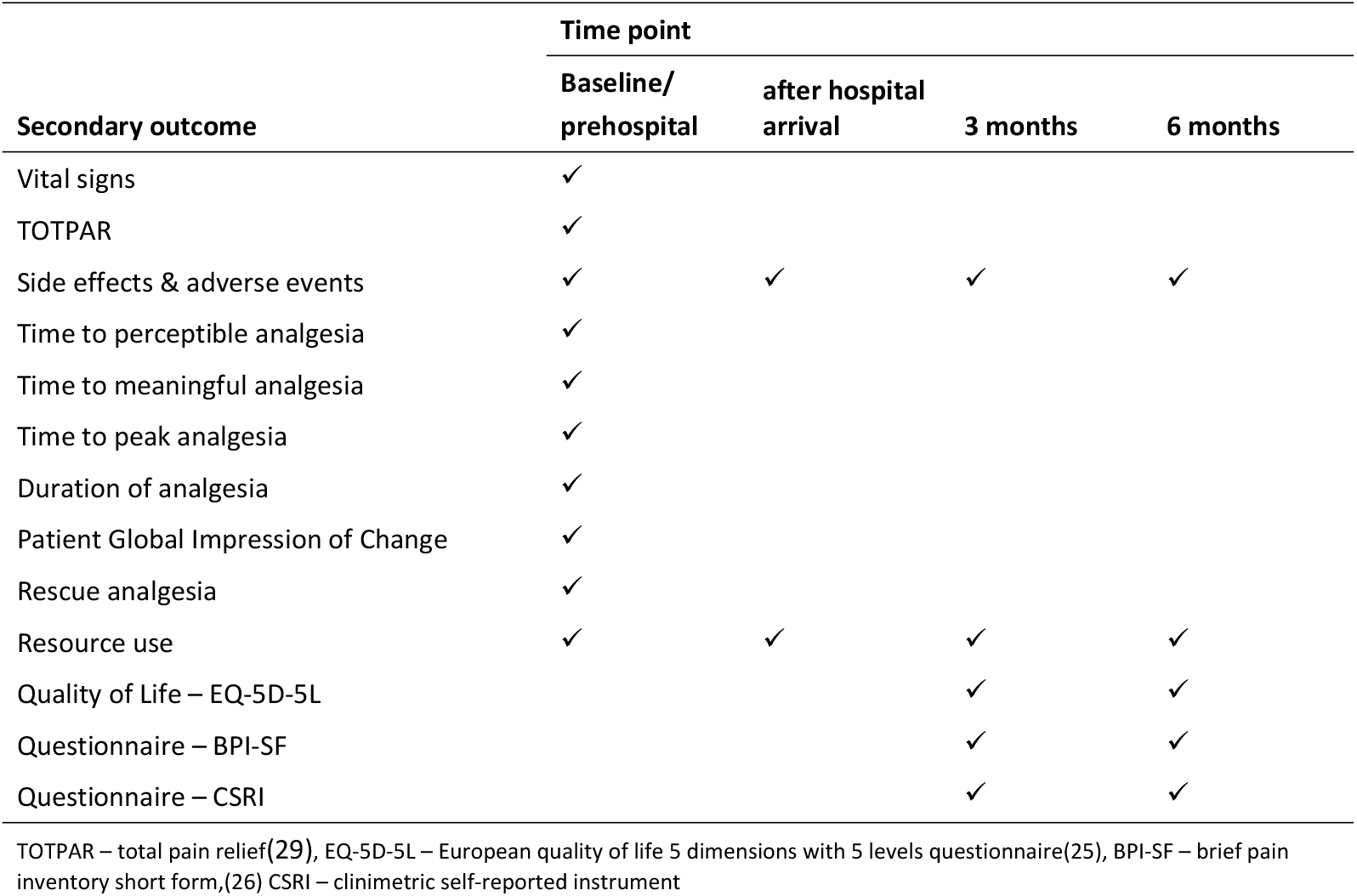
PACKMaN secondary outcomes and timepoints for measurement.

| Secondary outcome | Time point |  |  |  |
| --- | --- | --- | --- | --- |
|  | Baseline/<br>prehospital | after hospital<br>arrival | 3 months | 6 months |
| Vital signs | ✓ |  |  |  |
| TOTPAR | ✓ |  |  |  |
| Side effects & adverse events | ✓ | ✓ | ✓ | ✓ |
| Time to perceptible analgesia | ✓ |  |  |  |
| Time to meaningful analgesia | ✓ |  |  |  |
| Time to peak analgesia | ✓ |  |  |  |
| Duration of analgesia | ✓ |  |  |  |
| Patient Global Impression of Change | ✓ |  |  |  |
| Rescue analgesia | ✓ |  |  |  |
| Resource use | ✓ | ✓ | ✓ | ✓ |
| Quality of Life – EQ-5D-5L |  |  | ✓ | ✓ |
| Questionnaire – BPI-SF |  |  | ✓ | ✓ |
| Questionnaire – CSRI |  |  | ✓ | ✓ |

TOTPAR – total pain relief(29), EQ-5D-5L – European quality of life 5 dimensions with 5 levels questionnaire(25), BPI-SF – brief pain inventory short form,(26) CSRI – clinimetric self-reported instrument

## Statistical analysis

PACKMaN was powered to detect a 1-point difference in SPID score between morphine and ketamine using the 0-10 NRS. The sample size estimate assumed a standard deviation of 3.0,(31–33), a power of 90%, significance level of 5% and a withdrawal/non-response rate of 15%. (34–36) We calculated the trial required a sample of 446 participants, recruiting 223 to each arm of the study

In this secondary analysis, we dichotomise patients (irrespective of which trial drug they received) into two groups - those who reported a NRS pain score of below 4/10 on arrival at hospital, and those who reported a NRS pain score of 4/10 or higher, on arrival at hospital. If a participant did not have a pain score documented at hospital arrival, we used the last documented pain score prior to hospital arrival.(24) Consequently, our analysis compares outcomes for those patients whose pain severity was reduced from severe (NRS ≥ 7/10) to mild (NRS < 4/10), versus those who were still experiencing moderate or severe pain (NRS ≥ 4/10) on arrival at hospital. All analysis was undertaken using Stata SE v18.0. The primary outcome of this secondary analysis compares chronic pain outcomes, as defined by the Brief Pain Inventory short form (BPI_SF) at 3- and 6-months. Patient baseline demographic and injury characteristics were summarised across the two groupings: final NRS pain score < 4/10 and final NRS pain score ≥ 4/10. Continuous outcomes were summarised with means and standard deviations or medians and interquartile range and compared using a t-test, and categorical outcomes were summarised with frequencies and percentages using a chi-square test, in the case where an observation had fewer than 5 occurrences Fisher’s exact test was used.

Primary and secondary outcomes were summarised and analysed using unadjusted and adjusted regression models. The adjusted analysis was the primary analysis. The adjustment variables were chosen based on previous clinical research, where age and gender have been shown to have an impact on response to pain.(25, 26) In our explanatory analysis we identified a significant difference across final pain score category among participants who suffered injuries in their neck, pelvis, and lower limbs, as well as participant weight, for which we also performed adjusted analyses. Age was split into two categories participants aged below 65 years and those aged 65 years or above, weight was split into three categories determined by the 33.33% and 66.67% percentile.

Linear regression models were used for continuous outcomes and logistical regression models were used on binary outcomes. For categorical outcomes, ordered logistic regression models were used, and Cox’s proportional hazard model was employed to analyse time to event outcomes and duration of analgesia. Differences across the two groups and their 95% confidence intervals are presented alongside p-values.

## Results

PACKMaN recruited its first patient on 10/11/2021 and achieved its recruitment target on 31/05/2024. Patients were followed up for 6-months post-randomisation. Of the 449 participants randomised, 10 (2%) did not have a NRS pain score documented at hospital arrival, we imputed seven scores using the last documented pain score leaving 446 (99.3%) participants for analysis. 175 (39.2%) participants had a final NRS pain score below 4/10 and 271 (60.8%) had a final NRS pain score 4/10 or above. A CONSORT diagram details each group and rates of loss to follow up (Figure 1). Table 2 reports baseline characteristics between the groups.

**Table 2.** Participant characteristics by whether their final pain score was below 4 or not.

|  | Final pain<br>score below 4<br>(n=175) | Final pain<br>score 4 or<br>above<br>(n=271) | Total (n=446) | P-value |
| --- | --- | --- | --- | --- |
| Trial arm, n (%) |  |  |  |  |
| Arm A | 86 (49.1) | 142 (52.4) | 228 (51.1) |  |
| Arm B | 89 (50.9) | 129 (47.6) | 218 (48.9) | 0.50 |
| Age (years), mean (sd) | 66.28 (21.19) | 61.87 (21.93) | 63.60 (21.72) | 0.04 |
| Gender, n (%) |  |  |  |  |
| Female | 94 (53.7) | 144 (53.1) | 238 (53.4) |  |
| Male | 81 (46.3) | 127 (46.9) | 208 (46.6) | 0.91 |
| Ethnicity, n (%) |  |  |  |  |
| White | 84 (48.0) | 122 (45.0) | 206 (46.3) |  |
| Asian | 0 (0.0) | 1 (0.4) | 1 (0.2) |  |
| Any other ethnic group | 1 (0.6) | 1 (0.4) | 2 (0.4) |  |
| Ethnicity not given | 90 (51.4) | 147 (54.2) | 236 (53.0) | 0.89* |
| Weight, mean (sd) | 72.80 (17.07) | 78.73 (18.55) | 76.41 (18.2) | <0.001 |
| Initial pain score, mean (sd) | 8.76 (1.34) | 8.86 (1.21) | 8.82 (1.26) | 0.40 |
| Mechanism of injury, n (%) |  |  |  |  |
| Blunt trauma | 174 (99.4) | 266 (98.2) | 440 (98.7) |  |
| Penetrating trauma | 1 (0.6) | 4 (1.5) | 5 (1.1) |  |
| Burn | 0 (0.0) | 1 (0.4) | 1 (0.2) | 0.79* |
| Fracture/dislocation, n (%) |  |  |  |  |
| No | 14 (8.0) | 32 (11.8) | 46 (10.3) |  |
| Yes | 161 (92.0) | 239 (88.2) | 400 (89.7) | 0.20 |
| Soft tissue injury, n (%) |  |  |  |  |
| No | 125 (71.4) | 183 (67.5) | 308 (69.1) |  |
| Yes | 50 (28.6) | 88 (32.5) | 138 (30.9) | 0.38 |
| Wound/laceration, n (%) |  |  |  |  |
| No | 154 (88.0) | 235 (86.7) | 389 (87.2) |  |
| Yes | 21 (12.0) | 36 (13.3) | 57 (12.8) | 0.69 |
| Head, n (%) |  |  |  |  |
| No | 161 (92.0) | 249 (91.9) | 410 (91.9) |  |
| Yes | 14 (8.0) | 22 (8.1) | 36 (8.1) | 0.96 |
| Neck, n (%) |  |  |  |  |
| No | 171 (97.7) | 252 (93.0) | 423 (94.8) |  |
| Yes | 4 (2.3) | 19 (7.0) | 23 (5.2) | 0.03* |
| Chest, n (%) |  |  |  |  |
| No | 149 (85.1) | 211 (77.9) | 360 (80.7) |  |
| Yes | 26 (14.9) | 60 (22.1) | 86 (19.3) | 0.06 |
| Abdomen, n (%) |  |  |  |  |
| No | 173 (98.9) | 261 (96.3) | 434 (97.3) |  |
| Yes | 2 (1.1) | 10 (3.7) | 12 (2.7) | 0.14* |
| Pelvis, n (%) |  |  |  |  |
| No | 115 (65.7) | 202 (74.5) | 317 (71.1) |  |
| Yes | 60 (34.3) | 69 (25.5) | 129 (28.9) | 0.04 |
| Upper limbs, n (%) |  |  |  |  |
| No | 134 (76.6) | 209 (77.1) | 343 (76.9) |  |
| Yes | 41 (23.4) | 62 (22.9) | 103 (23.1) | 0.89 |
| Lower limbs, n (%) |  |  |  |  |
| No | 47 (26.9) | 98 (36.2) | 145 (32.5) |  |
| Yes | 128 (73.1) | 173 (63.8) | 301 (67.5) | 0.04 |

\*Fisher's exact test used for comparison

**Figure 1:**
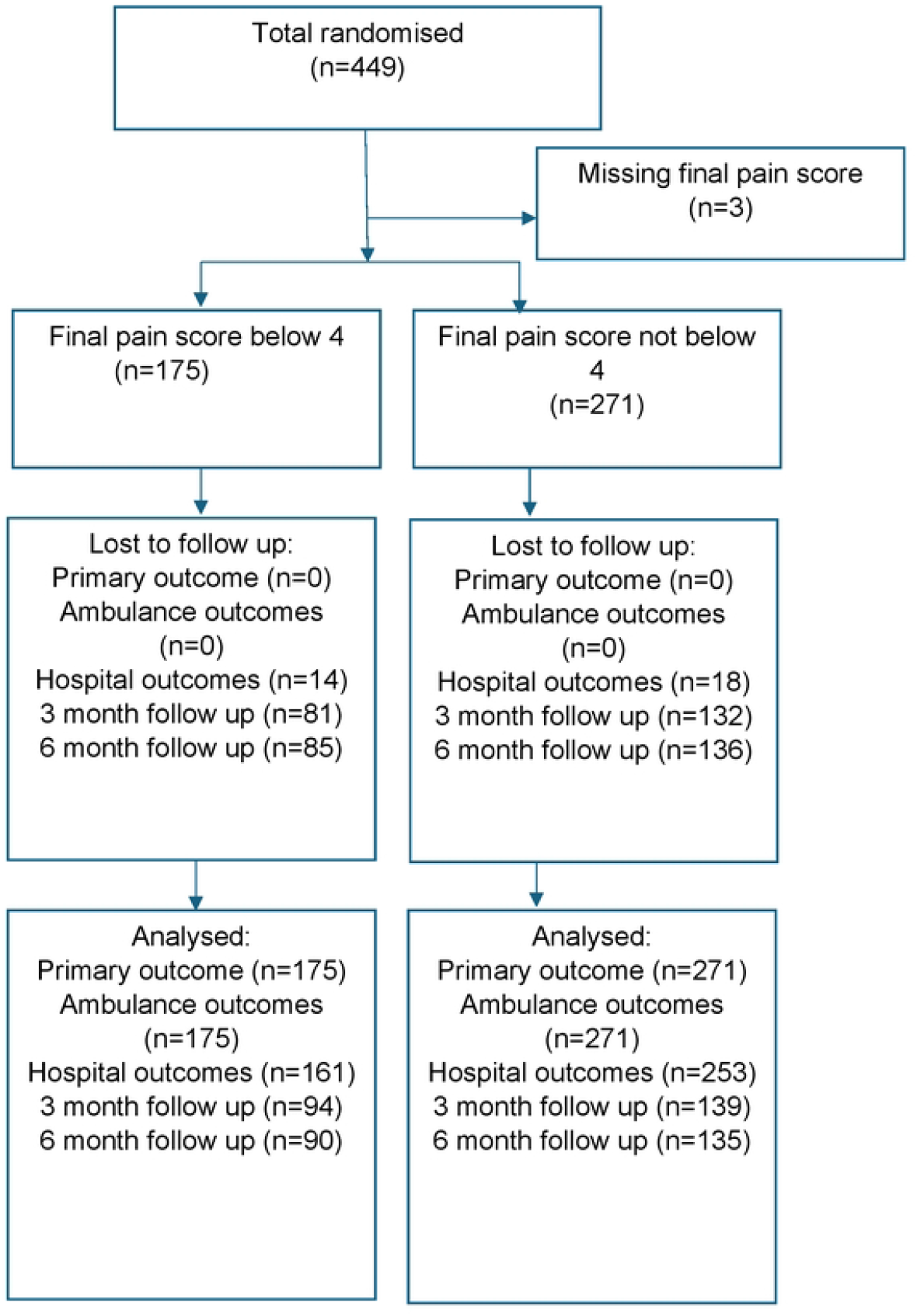
Allocation and outcomes dichotomised by final NRS score (<4 versus ≥ 4)

There were some minor differences in baseline characteristics between the two groups. Participants with a final NRS pain score < 4/10 had a mean age of 66.28 years compared to 61.87 years for participants with a NRS pain score ≥ 4/10 (p=0.04). The mean weight of participants with final NRS pain score < 4/10 was 72.80kg and 78.73kg for participants with final NRS pain score ≥ 4/10 (p<0.001). Participants with an injury to the neck were more likely to have a final NRS pain score ≥ 4/10 (p=0.03), whereas participants with an injury to the pelvis or lower limbs were more likely to have a NRS pain score < 4/10 (p=0.04).

For participants with a final NRS pain score < 4/10, the mean SPID score was 5.45 (SD=3.11) compared to 2.18 (SD=1.85) in those participants with a final NRS pain score ≥ 4/10 (MD, 3.24, 95% CI 2.78 to 3.70, p value<0.001, Table 3). Total Pain Relief (TOTPAR), global impression of change, time to perceptible analgesia, time to meaningful analgesia, and duration of analgesia (Table 3) also showed significant differences between the two participant groups. Patients with a final NRS pain score < 4/10 achieved more analgesia, as displayed by higher SPID and TOTPAR scores and global impression of change, while they also achieved pain relief quicker and for a longer duration of time. All these outcomes, as well as percentage of maximum dose in milligrams were also significant for the unadjusted analysis. Participants with a final NRS pain score < 4/10 received a lower percentage of maximum dose in milligrams but not when adjusted for our covariates, however there was no difference between dose in milligrams per kilogram. Time from randomisation to hospital arrival, ambulance job cycle time, and time to peak analgesia showed no difference across the two categories.

**Table 3.** Primary and Secondary outcomes summarised by final pain score status.

|  | Final pain score<br>below 4<br>(n=176) | Final pain<br>score 4 or<br>above<br>(n=271) | Unadjusted<br>difference<br>(95% CI); p-value | Adjusted<br>difference<br>(95% CI); p-value |
| --- | --- | --- | --- | --- |
| SPID, mean (sd) | 5.45 (3.11) | 2.18 (1.85) | - | MD, 3.24 (2.78 to 3.70; p<0.001) |
| TOTPAR, mean (sd) | 2.23 (1.09)<br>(n=172) | 0.83 (0.81)<br>(n=270) | MD, 1.40 (1.22 to 1.58); <0.001 | MD, 1.40 (1.22 to 1.57); <0.001 |
| Percentage of maximum dose in mg, mean (sd) | 0.58 (0.24)<br>(n=173) | 0.63 (0.26)<br>(n=270) | MD, -0.05 ( -0.10 to -0.00); 0.032 | MD, -0.05 (-0.10 to 0.00); 0.055 |
| Dose in mg per kilogram, mean (sd) | 0.21 (0.12) | 0.20 (0.10) | 0.01 (-0.01 to 0.03); 0.525 | -0.01 (-0.02 to 0.01); 0.579 |
| Time from randomisation to hospital arrival, mean (sd) | 52.58 (24.56)<br>(n=175) | 49.93 (26.04)<br>(n=271) | MD, 2.64 (-2.21 to 7.50); 0.285 | MD, 2.24 (-2.61 to 7.08); 0.365 |
| Ambulance job cycle time (minutes), mean (sd) | 86.97 (25.00)<br>(n=175) | 85.04 (27.23)<br>(n=271) | MD, 1.93 (-3.09 to 6.96); 0.45 | MD, 1.41 (-3.58 to 6.39); 0.579 |
| Time to perceptible analgesia, mean (sd) | 6 (3 - 10)<br>(n=171) | 8 (4 - 18)<br>(n=232) | HR, 1.89 (1.54 to 2.31); <0.001 | HR, 1.95 (1.58 to 2.39); <0.001 |
| Time to meaningful analgesia, mean (sd) | 12 (6 - 25)<br>(n=171) | 15 (7 - 33)<br>(n=131) | HR, 3.96 (3.12 to 5.02); <0.001 | HR, 4.19 (3.28 to 5.35); <0.001 |
| Time to peak analgesia, mean (sd) | 29.5 (10 - 48)<br>(n=172) | 19 (8 - 37.5)<br>(n=249) | HR, 0.97 (0.80 to 1.18); 0.769 | HR, 0.97 (0.79 to 1.18); 0.751 |
| Duration of analgesia, median (IQR) | 44.5 (31 - 56)<br>(n=172) | 34 (21 - 53)<br>(n=270) | HR, 0.77 (0.64 to 0.94); 0.009 | HR, 0.78 (0.64 to 0.95); 0.014 |
| Patient global impression of change, n(%) |  |  | OR, 0.06 (0.04 to 0.10); <0.001 | OR, 0.06 (0.04 to 0.10); <0.001 |
| Very much improved | 100 (57.5) | 26 (9.8) |  |  |
| Much improved | 67 (38.5) | 98 (36.8) |  |  |
| Minimally improved | 6 (3.4) | 107 (40.2) |  |  |
| No change | 0 (0.0) | 33 (12.4) |  |  |
| Minimally worse | 0 (0.0) | 1 (0.4) |  |  |
| Much worse | 0 (0.0) | 1 (0.4) |  |  |
| Very much worse | 1 (0.6) | 0 (0.0) |  |  |

Table 4 summarises patient in-hospital outcomes across the two NRS pain score categories. The outcome with a significant result in the adjusted analysis was patients admitted to hospital, where participants with a final NRS pain score < 4/10 were less likely to be admitted to hospital than the group with final NRS pain score ≥ 4/10 (OR, 0.37, 95% CI 0.21 to 0.65, p<0.001, Table 4). Ninety-seven (64.7%) patients with final NRS score < 4/10 were admitted to hospital compared to 163 (70.9%) in the group of patients with final NRS score ≥ 4/10. There was no difference in the unadjusted analysis. Number of days in critical care showed a significant difference of 6.25 days, with patients with a final NRS score < 4/10 spending more days in critical care. However, only 5 participants were admitted to critical care, 4 of whom had final NRS pain score ≥ 4/10 and 1 with final NRS pain score < 4/10. This finding should therefore be interpreted with caution. The small number of data points also meant the adjusted analysis related to days in critical care could not be completed.

**Table 4.** Hospital outcomes summarised by final pain score status.

|  | Final pain score<br>below 4 (n=176) | Final pain<br>score 4 or<br>above<br>(n=271) | Unadjusted<br>difference<br>(95% CI); p-value | Adjusted<br>difference<br>(95% CI); p-value |
| --- | --- | --- | --- | --- |
| Length of stay in ED<br>(minutes), mean (sd) | 565.75 (359.84)<br>(n=149) | 567.50<br>(367.74)<br>(n=229) | MD, -1.75 (-77.21<br>to 73.72); 0.964 | MD, -9.68 (-87.18<br>to 67.82); 0.806 |
| Length of stay in hospital<br>(days), mean (sd) | 11.00 (54.60)<br>(n=88) | 14.05 (31.24)<br>(n=156) | MD, -3.05 (-13.87<br>to 7.76); 0.579 | MD, -7.95 (-19.02<br>to 3.12); 0.158 |
| Admitted to hospital, n(%) |  |  |  |  |
| No | 53 (35.3) | 67 (29.1) | OR, 0.75 (0.49 to<br>1.17); 0.204 | OR, 0.37 (0.21 to<br>0.65); <0.001 |
| Yes | 97 (64.7) | 163 (70.9) |  |  |
| Admitted to critical care,<br>n(%) |  |  |  |  |
| No | 96 (99.0) | 159 (97.5) | OR, 0.41 (0.05 to<br>3.76); 0.433 | OR, 0.66 (0.04 to<br>9.85); 0.767 |
| Yes | 1 (1.0) | 4 (2.5) |  |  |
| Participant had a CT scan,<br>n(%) |  |  |  |  |
| No | 100 (66.7) | 140 (61.1) | OR, 0.79 (0.51 to<br>1.21); 0.275 | OR, 0.93 (0.59 to<br>1.49); 0.776 |
| Yes | 50 (33.3) | 89 (38.9) |  |  |
| Number of CT scans, mean<br>(sd) | 2.04 (2.14)<br>(n=50) | 2.88 (2.66)<br>(n=89) | MD, -0.84 (-1.71<br>to .03); 0.059 | MD, -0.41 (-1.24<br>to 0.43); 0.336 |
| Days in critical care, mean<br>(sd) | 10.00 (.)<br>(n=1) | 3.75 (1.26)<br>(n=4) | MD, 6.25 (1.77 to<br>10.7); 0.021 | N/A |
| Number of paramedics,<br>mean (sd) | 1.27 (0.58)<br>(n=175) | 1.25 (0.48)<br>(n=271) | MD, 0.02 (-0.08<br>to 0.12); 0.697 | MD, 0.03 (-0.08<br>to 0.13); 0.617 |
| Number of technicians,<br>mean (sd) | 0.57 (0.66)<br>(n=175) | 0.53 (0.58)<br>(n=271) | MD, 0.04 (-0.08<br>to 0.15); 0.52 | MD, 0.01 (-0.11<br>to 0.12); 0.897 |
| Number of doctors, mean<br>(sd) | 0.02 (0.13)<br>(n=175) | 0.00 (0.06)<br>(n=271) | MD, 0.01 (-0.01<br>to 0.03); 0.142 | MD, 0.02 (-0.00<br>to 0.03); 0.102 |
| Number of other, mean<br>(sd) | 0.41 (0.50)<br>(n=174) | 0.41 (0.53)<br>(n=267) | MD, -0 (-0.10 to<br>0.10); 0.997 | MD, 0.02 (-0.08<br>to 0.12); 0.737 |

Responses to the brief pain inventory short form (BPI-SF) at 3-months post randomisation are presented in table 5. Four outcomes showed a significant difference in the adjusted analysis across final NRS score pain intensity at worst (MD, −1.09, 95% CI −1.90 to −0.28, p=0.008), pain intensity at its least (MD, −0.70, 95% CI −1.32 to −0.09, p=0.024), pain intensity on average (MD, −0.76, 95% CI −0.1.41 to −0.11, p=0.022) and pain intensity right now (MD, −0.88, 95% CI −1.62 to −0.14, p=0.021). These results suggest that participants with a final NRS pain score < 4/10 experienced less pain at all levels 3-months post injury. Looking at the pain summary outcomes from the brief pain inventory pain severity score (MD, −0.90, 95% CI −1.54 to −0.27, p=0.006) and pain inference score (MD, −1.15, 95% CI −1.91 to −0.38, p=0.003), showing overall less chronic pain is experienced by patients with a final NRS pain score < 4/10. All outcomes from the brief pain inventory except for pain intensity on average showed a significant difference for the unadjusted analysis also.

**Table 5.** Brief pain inventory outcomes at 3-months by final pain score status.

| Pain category | Final pain score<br>below 4<br>(n=176) | Final pain<br>score 4 or<br>above<br>(n=271) | Unadjusted<br>difference<br>(95% CI); p-value | Adjusted<br>difference<br>(95% CI); p-value |
| --- | --- | --- | --- | --- |
| Pain rate at worst<br>(scaled 0-10), mean (sd) | 5.21 (3.10)<br>(n=90) | 6.12 (2.83)<br>(n=138) | MD, -0.91 ( -1.70<br>to -0.13); 0.023 | MD, -1.09 (-1.90<br>to -0.28); 0.008 |
| Pain rate at least<br>(scaled 0-10), mean (sd) | 2.60 (2.10)<br>(n=89) | 3.20 (2.28)<br>(n=135) | MD, -0.60 ( -1.20<br>to -0.01); 0.046 | MD, -0.70 (-1.32<br>to -0.09); 0.024 |
| Pain rate at average<br>(scaled 0-10), mean (sd) | 4.17 (2.40)<br>(n=89) | 4.77 (2.33)<br>(n=133) | MD, -0.61 (-1.24<br>to 0.03); 0.062 | MD, -0.76 (-1.41<br>to -0.11); 0.022 |
| Pain rate right now<br>(scaled 0-10), mean (sd) | 3.30 (2.70)<br>(n=90) | 4.13 (2.76)<br>(n=133) | MD, -0.83 ( -1.56<br>to -0.09); 0.028 | MD, -0.88 (-1.62<br>to -0.14); 0.021 |
| Severity score (<br>scaled 0-10), mean (sd) | 3.84 (2.36)<br>(n=90) | 4.62 (2.30)<br>(n=138) | MD, -0.78 ( -1.40<br>to -0.16); 0.014 | MD, -0.90 (-1.54<br>to -0.27); 0.006 |
| Pain inference score<br>(scaled 0-10), mean (sd) | 4.08 (2.63)<br>(n=89) | 5.07 (2.99)<br>(n=137) | MD, -0.99 ( -1.76<br>to -0.22); 0.012 | MD, -1.15 (-1.91<br>to -0.38); 0.003 |

Table 6 summarises participant responses to the brief pain inventory short form at 6-months post randomisation. The following outcomes show a significant difference in the adjusted analysis across final NRS score: pain intensity at worst (MD, −1.17, 95% CI −2.07 to −0.27, p=0.011), pain intensity on average (MD, −0.89, 95% CI −1.65 to −0.13, p=0.021) and current pain intensity (MD, −0.78, 95% CI −1.53 to −0.02, p=0.043). As in the 3-month follow up, these results also suggest that participants with a final NRS pain score < 4/10 experience less pain at all levels, except for pain intensity at its least, 6-months post injury, pain severity score (MD, −0.82, 95% CI −1.52 to −0.12, p=0.022) and pain inference score (MD, −1.1, 95% CI −1.9 to −0.29, p=0.008) showed significant differences in chronic pain at 6-months post injury, where patients who had a final NRS pain score < 4/10 experienced fewer chronic pain issues. All these outcomes also showed a significant difference in the unadjusted analysis.

**Table 6.** Brief pain inventory outcomes at 6-months by final pain score status.

| **Pain category** | **Final pain score**<br>**below 4**<br>**(n=176)** | **Final pain**<br>**score 4 or**<br>**above**<br>**(n=271)** | **Unadjusted**<br>**difference**<br>**(95% CI); p-value** | **Adjusted**<br>**difference**<br>**(95% CI); p-value** |
| --- | --- | --- | --- | --- |
| Pain rate at worst<br>(scaled 0-10), mean (sd) | 4.59 (3.28)<br>(n=85) | 5.78 (3.11)<br>(n=133) | MD, -1.19 ( -2.06<br>to -0.32); 0.007 | MD, -1.17 (-2.07<br>to -0.27); 0.011 |
| Pain rate at least<br>(scaled 0-10), mean (sd) | 2.58 (2.26)<br>(n=85) | 3.12 (2.45)<br>(n=133) | MD, -0.54 (-1.20<br>to 0.11); 0.101 | MD, -0.45 (-1.13<br>to 0.23); 0.191 |
| Pain rate at average<br>(scaled 0-10), mean (sd) | 3.80 (2.67)<br>(n=85) | 4.66 (2.69)<br>(n=131) | MD, -0.86 ( -1.59<br>to -0.12); 0.023 | MD, -0.89 (-1.65<br>to -0.13); 0.021 |
| Pain rate right now<br>(scaled 0-10), mean (sd) | 2.96 (2.52)<br>(n=85) | 3.72 (2.75)<br>(n=133) | MD, -0.76 ( -1.49<br>to -0.03); 0.042 | MD, -0.78 (-1.53<br>to -0.02); 0.043 |
| Severity score<br>(scaled 0-10), mean (sd) | 3.48 (2.43)<br>(n=85) | 4.32 (2.51)<br>(n=133) | MD, -0.84 ( -1.52<br>to -0.16); 0.016 | MD, -0.82 (-1.52<br>to -0.12); 0.022 |
| Pain inference score<br>(scaled 0-10), mean (sd) | 3.44 (2.60)<br>(n=85) | 4.51 (3.07)<br>(n=132) | MD, -1.07 ( -1.86<br>to -0.28); 0.008 | MD, -1.10 (-1.90<br>to -0.29); 0.008 |

## Discussion

In this analysis we found that participants who achieved a final NRS pain score < 4/10 sustained greater pain relief, defined by SPID, compared to participants who reported a final NRS pain score ≥ 4/10. These findings are not unexpected given that a lower final pain score at hospital arrival is likely to be associated with a greater reduction in pain intensity throughout the ambulance journey, and thus greater pain relief. We have ruled out a potential link between pain management and prehospital assessment time period, showing there is no difference between final NRS pain score and the period of time one is assessed (time from randomisation to hospital arrival and ambulance job cycle time). It is important to have ruled out this potential link as it means there is less risk of bias being incurred due to the length of ambulance journey between both groups.

We found no difference in the unadjusted analysis of participants admitted to hospital between the two groups; however, the opposite was found in the adjusted analysis of some outcomes. This may be due to the inclusion of age as a covariate in the adjusted analysis. Age is associated with final pain score grouping and we have found that whether a participant was admitted to hospital is also dependent on age, which could contribute to this finding.

Although the percentage of total dose in milligrams showed a significant difference in the unadjusted analysis, it was no longer significant in the adjusted analysis. The significant result in the unadjusted analysis suggests that patients with a final pain NRS score < 4/10 received a smaller dose than those who had a final NRS pain score ≥ 4/10. One plausible explanation is that those with a lower and more stable pain score would require less investigational medicinal product (IMP) to manage their pain. However, we have also shown that participants who did not achieve a final NRS pain score < 4/10 were heavier than those who did, and paramedics are advised to give weight dependent doses of IMP, where heavier patients receive a higher dose. This would be a stronger explanation as to why participants with a final pain score ≥ 4/10 received a higher dose, due to the confounding variable weight. When considering participant doses in milligrams per kilogram, we found no difference across the final pain score groups, supporting this explanation.

Our findings for the long-term outcomes of the BPI-SF questionnaire build on what has been reported in previous studies and adds some important novel context. Rivara et al(37) found that participants admitted to hospital due to acute trauma still experience moderately severe pain from their injuries at one-year. In that study, adult participants with at least one injury and an Abbreviated Injury Scale (AIS) score of 3 or higher were assessed using the Chronic Pain Grade Scale 12-months post injury.(37) The study included 3047 patients, of whom 63% still reported pain 12-months post injury. Williamson et al(38) conducted a prospective cohort study of adults with orthopaedic injuries and found that 30% of all participants had moderate or severe pain 6-months after injury, where severe or moderate pain at discharge was found to be a predictor of this. Daoust et al(39) conducted a retrospective observational cohort study of adult patients admitted for injury in trauma centres 3 to 12 months post injury, where 15% patients developed chronic pain. Daoust found the following factors associated with chronic pain, having a spinal cord injury, disc-vertebra trauma, history of alcoholism, anxiety, and depression, and being female.(39) In our analysis we have found a similar conclusion, demonstrating a relationship between prehospital management of acute pain and chronic pain at 3- and 6-months post injury. Our study has built on previous findings as our analysis population included only participants experiencing severe pain and suggests that for patients reporting severe pain following acute traumatic injury, failure to provide sufficient pre-hospital analgesia to reduce pain intensity to mild levels (NRS<4) on hospital arrival is associated with increased risk of chronic pain. We believe these findings will be important to a wide range of EMS systems where paramedics provide parenteral analgesia for patients with severe pain following trauma.

This analysis is not without limitations. The primary aim of the PACKMAN trial was to compare effectiveness of ketamine and morphine; therefore, our conclusions should be interpreted with caution. Some confounding variables such as age and weight were not balanced across final pain score groups and this may have had an impact on our adjusted analyses.

It is also plausible that there is a direct relationship between injury severity, and both analgesic efficacy and chronic pain. The PACKMaN study did plan to explore the relationship between injury severity, analgesic efficacy and longer-term outcomes employing data from the Trauma and Audit Research Network (TARN) Database as a proxy for injury severity. Unfortunately, a cyberattack on the NHS infrastructure in 2023 severely disrupted reporting to the TARN database. Consequently, we were unable to obtain this data for a significant proportion of participants and therefore unable to reliably perform any sensitivity analysis related to injury severity. It is possible that injury severity is the causative factor behind both failures to achieve a NRS pain score < 4/10 on hospital arrival and the likelihood of experiencing chronic pain. Failure to achieve a NRS pain score < 4/10 on hospital arrival may simply be correlated to the risk of chronic pain, rather than a causative factor.

Finally, the PACKMAN trial experienced a drop-off in participants completing follow up, which could introduce bias to the brief pain inventory results as participants with more pain at follow-up may be more or less likely to complete follow-up.

## Conclusion

In this secondary analysis of data from the PACKMaN trial we were able to demonstrate a statistically significant relationship between failure to achieve a reduction in pre-hospital NRS pain score from severe (≥ 7/10) to mild (< 4/10) on arrival at hospital and the risk of chronic pain at both 3- and 6-months. The limitations of utilising data in secondary analysis highlights an urgent need for further research to identify how prehospital pain management may impact longer-term patient outcomes.

## Data Availability

Data from the PACKMaN study will be available upon request from the Warwick Clinical Trials Unit (WCTU) Data Sharing Committee 6 months after the publication of the funder report. All applications will be assessed by according to WCTU Standard Operating Procedures available at: https://warwick.ac.uk/fac/sci/med/research/ctu/ctuintranet/qa/sop/sop153_sharingdata_v4.0.pdf. An application form to access data can be obtained from, which should be submitted to the corresponding author in the first instance. Any data shared will be anonymised - no identifiable data will be available. There is no fixed end-date for data availability.

## Acknowledgements

We are indebted to our PPI colleague, Mr Duncan Buckley, who provided input into the original design of the trial including our approach to consent and the outcomes measured. We acknowledge the outstanding contribution of research teams at WMAS (Andrew Rosser, Imogen Gunson, Joshua Miller, Owen Stanley, Zoe Green, Christine Evans, Paul Lockley, Brian Davidson, Rhiannon Boldy and Robert Millard) and YAS (Fiona Bell, Elisha Miller, Richard Pilbery, Alexander Diffley, Kelly Hird, Andrew Pountney, Julian Mark, Sean Jennings, Daniel Lindley, Rachel Byrne and Cath Hodson). We are extremely grateful to all the paramedics and patients who participated in the PACKMaN trial. We thank the members of the Clinical Research Ambassadors Group who helped to design patient facing documentation. Finally, we thank the National Institute of Health and Care Research (NIHR) Health Technology Assessment Programme who funded the research (128086).

